# Clinical outcomes after the introduction of dolutegravir for second-line antiretroviral therapy in South Africa: a retrospective cohort study

**DOI:** 10.1101/2023.07.07.23292347

**Authors:** Kwabena Asare, Yukteshwar Sookrajh, Johan van der Molen, Thokozani Khubone, Lara Lewis, Richard J Lessells, Kogieleum Naidoo, Phelelani Sosibo, Rosemary van Heerden, Nigel Garrett, Jienchi Dorward

## Abstract

**Background:** Dolutegravir is now recommended for second-line anti-retroviral therapy (ART) in low- and middle-income countries. We compared outcomes with dolutegravir (DTG) versus the previous lopinavir/ritonavir (LPV/r) regimen in South Africa.

**Methods:** We used routinely collected, de-identified data from 59 South African clinics. We included people living with HIV aged ≥ 15 years with virologic failure (two consecutive viral loads ≥1000 copies/mL) on first-line tenofovir disoproxil fumarate (TDF)-based ART and switched to second-line ART. We used modified Poisson regression models to compare outcomes of 12-month retention-in-care and viral suppression (<50 copies/ml) after switching to second-line regimens of zidovudine (AZT), emtricitabine/lamivudine (XTC), DTG and TDF/XTC/DTG and AZT/XTC/LPV/r.

**Findings:** Of 1214 participants, 729 (60.0%) were female, median age was 36 years (interquartile range 30 to 42), 689 (56.8%) were switched to AZT/XTC/LPV/r, 217 (17.9%) to AZT/XTC/DTG and 308 (25.4%) to TDF/XTC/DTG. Retention-in-care was higher with AZT/XTC/DTG (85.7%, adjusted risk ratio (aRR) 1.14, 95% confidence interval (CI) 1.03 to 1.27; adjusted risk difference (aRD) 10.89%, 95%CI 2.01 to 19.78) but not different with TDF/XTC/DTG (76.9%, aRR 1.01, 95%CI 0.94 to 1.10; aRD 1.04%, 95%CI -5.03 to 7.12) compared to AZT/XTC/LPV/r (75.2%). Retention-in-care with TDF/XTC/DTG was not statistically significantly different from AZT/XTC/DTG (aRR 0.89, 95%CI 0.78 to 1.01; aRD - 9.85%, 95%CI -20.33 to 0.63). Of 799 participants who were retained-in-care with a 12-month viral load, viral suppression was higher with AZT/XTC/DTG (59.3%, aRR 1.25, 95%CI 1.06 to 1.47; aRD 11.57%, 95%CI 2.37 to 20.76) and TDF/XTC/DTG (60.7%, aRR 1.30, 95%CI 1.14 to 1.48; aRD 14.16%, 95%CI 7.14 to 21.18) than with the AZT/XTC/LPV/r regimen (46.7%).

**Interpretation:** DTG-based second-line regimens were associated with similar or better retention-in-care and better viral suppression than the LPV/r-based regimen. TDF/XTC/DTG had similar viral suppression compared to AZT/XTC/DTG.

**Funding:** Bill & Melinda Gates Foundation, Africa Oxford Initiative.

## RESEARCH IN CONTEXT

### Evidence before this study

We searched PubMed from inception until May 30, 2023, with no language restrictions, for published articles evaluating outcomes with dolutegravir-zidovudine-based versus dolutegravir-tenofovir disoproxil fumarate-based versus lopinavir-ritonavir-based regimens for second-line anti-retroviral therapy. We used search words [dolutegravir AND (tenofovir OR lopinavir-ritonavir) AND (second-line anti-retroviral therapy)]. We found 5 clinical trials (DAWNING, NADIA, D2EFT, VISEND and ARTIST) and zero observational studies. The DAWNING trial showed the superiority of dolutegravir versus ritonavir-boosted lopinavir, when used with two nucleoside reverse transcriptase inhibitors (NRTIs) in 624 participants with previous first-line failure (≥ 400 copies/ml) with non-nucleoside reverse transcriptase inhibitor (NNRTI) based regimens. At week 48 after baseline, 261 of 312 (84.0%) participants in the dolutegravir group achieved viral suppression (< 50 copies/ml) compared with 219 of 312 (70.0%) in the ritonavir-boosted lopinavir group. Among 464 participants in the NADIA trial with first-line treatment failure (≥ 1000 copies/ml) on an NNRTI with tenofovir and lamivudine or emtricitabine, recycled tenofovir for second-line treatment was non-inferior at week 48 compared to zidovudine (90.2% vs 91.7%) all used with dolutegravir or darunavir for viral suppression (< 400 copies/ml). The VISEND and D2EFT trials demonstrated the non-inferiority of dolutegravir with tenofovir and lamivudine or emtricitabine to standard-of-care ritonavir-boosted protease inhibitors lopinavir, atazanavir and darunavir for second-line treatment. In the single-arm ARTIST trial, including 62 participants with virologic failure on first-line tenofovir and lamivudine or emtricitabine with efavirenz or nevirapine and switched to second-line regimens with recycled tenofovir and dolutegravir, viral suppression (< 50 copies/ml) was 74.0% at 48 weeks. These clinical trials, except the ARTIST trial, have demonstrated the effectiveness of second-line DTG used with AZT or recycled first-line tenofovir for viral suppression compared to previous standard-of-care ritonavir-boosted protease inhibitor-based regimens. However, outcomes in non-trial or routine healthcare settings, where treatment adherence might be relatively lower than in trial settings, are scarce. Furthermore, the relative effectiveness of these second-line regimens on retention-in-care, probably due to regimen tolerability within an anti-retroviral treatment programme setting, is also limited.

### Added value of this study

After the implementation of dolutegravir for second-line anti-retroviral treatment in low- and middle-income countries, this is the first study using ART programme data from routine care clinics to assess outcomes after switching to second-line dolutegravir used with zidovudine or recycled first-line tenofovir versus the previously recommended ritonavir-boosted lopinavir on 12-month retention-in-care and viral suppression. Dolutegravir was better when used with zidovudine but was similar when used with recycled tenofovir for retention-in-care, and all were better for viral suppression versus the previous ritonavir-lopinavir-based regimen. Dolutegravir used with recycled tenofovir was slightly lower but not significantly different for retention-in-care and similar for viral suppression versus when used with the standard zidovudine.

### Implications of all the available evidence

Evidence from ongoing real-world cohorts through anti-retroviral treatment programmatic data evaluation is important for confirming the usefulness of common regimen combinations in regular healthcare settings to guide further decision-making. We have provided evidence outside clinical trial settings supporting WHO’s recommendation of dolutegravir use replacing lopinavir-ritonavir for second-line treatment in resource-limited settings. Our findings also suggest that recycling first-line tenofovir instead of replacing it with zidovudine for a dolutegravir-based second-line regimen can be an effective alternative for viral suppression. Further evidence from routine care settings on adverse events during second-line dolutegravir-based treatment would be a vital addition to evidence for continuous improvement of anti-retroviral treatment guidelines.

## INTRODUCTION

Following World Health Organization (WHO)^1, 2^ recommendations, dolutegravir (DTG) has been implemented for second-line antiretroviral therapy (ART) in South Africa since December 2019, replacing previously recommended regimens with lopinavir-ritonavir (LPV/r)^3, 4^. The WHO recommendation was based on results from the DAWNING trial^5^ showing superior efficacy of DTG for second-line ART compared to LPV/r. Furthermore, evidence from the NADIA^6^ trial suggested that recycling or maintaining first-line tenofovir disoproxil fumarate (TDF) in a DTG-based second-line ART was non-inferior to switching to Zidovudine (AZT). However, there is little data from routine care demonstrating the effectiveness of DTG, either with AZT or recycling TDF, on clinical outcomes during second-line ART.

Before December 2019, people living with HIV (PLHIV) in South Africa who were receiving the standard first-line regimen of TDF, emtricitabine (FTC) and efavirenz (EFV), and presented with virologic failure (repeat viral load ≥ 1000 copies/ml two to three months apart), were recommended to switch to second-line regimen of zidovudine (AZT), lamivudine (3TC) and LPV/r.^4^ After DTG was introduced for second-line ART in 2019, they were recommended to switch to AZT/3TC/DTG. Some people with virologic failure during first-line treatment may have been switched to TDF/3TC/DTG, either inadvertently as part of the transition to first-line dolutegravir or by clinicians following preliminary evidence suggesting that TDF/3TC/DTG may be an effective second-line regimen.^7^ As the rollout of DTG in low- and middle-income countries (LMICs) continues^8, 9^, evidence on the effectiveness of different regimens in routine care settings is required to guide further rollout and confirm clinical trial findings.^7^

Therefore, we aimed to evaluate the effectiveness of DTG plus emtricitabine/lamivudine (XTC) in combination with AZT or TDF versus the previously recommended regimen AZT/XTC/LPV/r for second-line treatment in routine care settings.

## METHODS

### Study design and setting

We did a retrospective cohort study with de-identified, routinely collected data from South Africa’s ART program in 59 primary healthcare facilities in the eThekwini Municipality of the KwaZulu-Natal province. South Africa’s ART delivery in public healthcare clinics involves clinical assessment for pregnancy, viral load, and CD4 count testing and screening for tuberculosis at baseline ART initiation and follow-up visits.^4^ Viral load is repeated at 6 and 12 months after ART initiation and 12-monthly thereafter. CD4 count is measured at ART initiation and 12 months thereafter and then only repeated if clinically indicated (e.g., viral load ≥ 1000 copies/ml). PLHIV with a viral load ≥1000 copies/ml are recommended to receive enhanced adherence counselling and a repeat viral load after two to three months. For people receiving first-line regimens containing a non-nucleoside reverse transcriptase inhibitor (NNRTI) such as efavirenz or nevirapine, virological failure is defined as two consecutive viral loads ≥ 1000 copies/ml two to three months apart and switching to second-line ART is recommended. There is no routine HIV drug resistance testing at the time of first-line ART failure in this setting. The study was approved by the Biomedical Research Ethics Committee of the University of Kwazulu-Natal (BE646/17), the KwaZulu-Natal Provincial Health Research Ethics Committee (KZ_201807_021), the TB/HIV Information Systems Data Request Committee, and the eThekwini Municipality Health Unit.

### Participants

Our study population included all PLHIV aged ≥ 15 who were switched to a second-line ART regimen between December 1, 2019, and November 30, 2020. We used this baseline period of second-line switching to allow a minimum of 12 months follow-up duration plus 180 days before the data cutoff on April 21, 2022. We excluded people not previously receiving standard first-line regimens of TDF/XTC/EFV or TDF/XTC/NVP at the time of virologic failure and those not switched to second-line regimens of AZT/XTC/LVP/r, AZT/XTC/DTG and TDF/XTC/DTG. Thus, we excluded people who were switched to a four-drug regimen of AZT/3TC/TDF plus LPV/r or DTG (i.e. hepatitis B coinfected participants) and those switched to abacavir-based regimens. We also excluded people who did not strictly meet guideline-defined first-line virologic failure criteria (two consecutive viral loads ≥ 1000 copies/ml at least 56 days apart).

### Data sources and data management

We used data from South Africa’s TIER.Net electronic database, which contains demographics, clinical status, regimen and clinic visit information of people receiving ART in public sector healthcare clinics.^10^ Data were de-identified by the South African National Department of Health’s TB/HIV Information Systems (THIS -www.tbhivinfosys.org.za/) before access and analysis by the study team.

### Outcomes

Our primary outcomes were retention-in-care and viral suppression at 12 months after starting second-line treatment. Retention-in-care at 12 months was defined as not being lost to follow-up or recorded in TIER.Net as either deceased or ‘transferred out’ to another clinic (as we could not access or link to data at other clinics to establish retention-in-care) by 365 days after starting second-line treatment. We defined loss to follow-up using the South African ART programme guidelines of being 90 days late for a visit^11^ and used the date of last visit as the date of loss-to-follow-up. Viral suppression was defined as viral load < 50 copies/ml. We included one secondary outcome for a post-hoc sensitivity analysis defining viral suppression as viral load < 1000 copies/ml. Because viral loads are not always completed regularly in routine care, we defined the 12-month window as the closest viral load to 365 days between 181 to 545 days after starting second-line treatment and included only the viral loads of participants retained in care.

### Exposures

The primary exposure was the second-line ART regimen combination (AZT/XTC/DTG or TDF/XTC/DTG or AZT/XTC/LPV/r) that participants were switched to after virologic failure. Secondary exposures included participant baseline characteristics when starting second-line treatment, such as age, gender, active tuberculosis, most recent viral load, most recent CD4 count, and time on ART.

### Statistical analyses

We performed all statistical analyses using R 4.2.0 (R Foundation for Statistical Computing, Vienna, Austria)^12^ and STATA 17.0^13^. We summarised participants’ baseline demographic, clinical characteristics, and outcomes at 12 months follow-up. We used percentages and medians to describe the baseline characteristics and assessed missing data stratified by the second-line regimen. We conducted univariable and multivariable modified Poisson regression with robust standard errors adjusting for clustering by clinic^14^ to determine the risk ratios of retention-in-care and viral suppression at 12 months follow-up. In all regression analyses, we compared the two DTG-based regimens of AZT/XTC/DTG and TDF/XTC/DTG versus AZT/XTC/LPV/r that participants were originally prescribed when starting second-line treatment and reported the risk differences. We also present results from these models comparing TDF/XTC/DTG versus AZT/XTC/DTG. We adjusted for participant characteristics at baseline, namely age category, gender, active tuberculosis, and category for recent viral load, in the multivariable regression models. We did not include the most recent CD4 count or time on ART in the multivariable models, as the resultant predicted probabilities exceeded one. Instead, we conducted sensitivity analyses of the effect of the ART regimen on each outcome, adjusted for only CD4 count and time on ART, to demonstrate a lack of confounding by these variables. We conducted further sensitivity analyses excluding participants who changed their originally prescribed second-line regimen within 12 months after baseline.

## RESULTS

From December 1, 2019, to November 30, 2020, 1672 people were recorded as switching to second-line ART after virologic failure (two consecutive viral loads ≥ 1000 copies/ml at least 56 days apart) while receiving first-line ART at the study clinics (Figure 1). We excluded 302 participants who were not previously receiving standard first-line regimens of TDF/XTC/EFV or TDF/XTC/NVP at the time of virologic failure and 156 who were not switched to standard second-line regimens of AZT/XTC/LPV/r or AZT/XTC/DTG or TDF/XTC/DTG. Of the remaining 1214 participants who were included in this analysis, 689 (56.8 %) were switched to AZT/XTC/LPV/r, 217 (17.9%) were switched to AZT/XTC/DTG, and 308 (25.4%) were switched to TDF/XTC/DTG second-line regimens.

**Figure 1.**
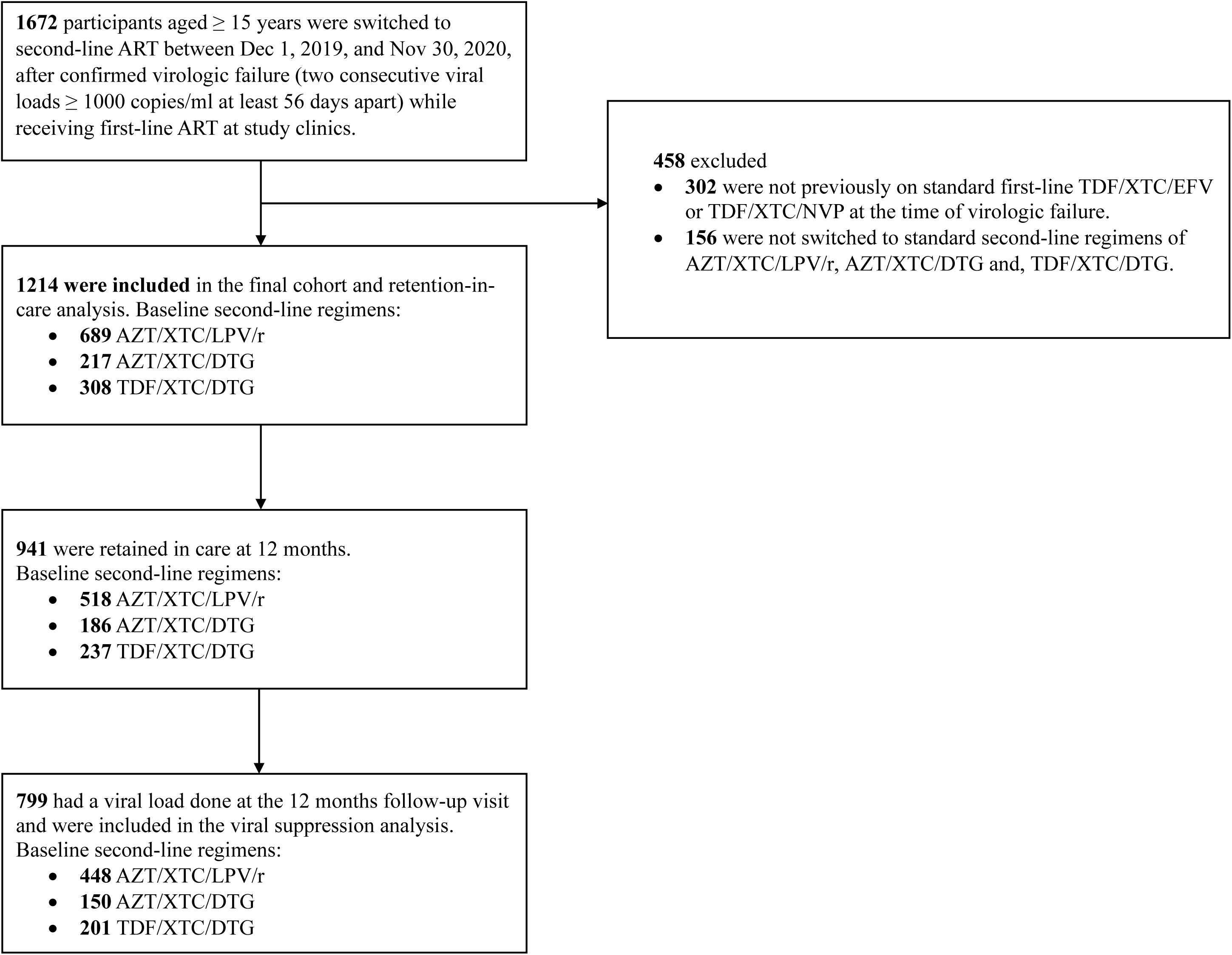
Flow diagram of participants receiving care at 59 clinics in South Africa. ART = Antiretroviral treatment, AZT = Zidovudine, DTG = Dolutegravir, EFV = Efavirenz, LVP/r = Lopinavir-ritonavir, ml = Milliliter, NVP = Nevirapine, PLHIV = People living with HIV, TDF = Tenofovir disoproxil fumarate, XTC = Emtricitabine or Lamivudine.

Overall, the median age was 36 (interquartile range (IQR) 30-42), and 60.0% (n = 729) were female (Table 1). Almost all participants previously received first-line TDF/XTC/EFV (n = 1198, 98.7%). Age was similar between the three regimen groups, but there were more females in the AZT/XTC/LPV/r group (n = 460, 66.8%) than in the AZT/XTC/DTG (n = 108, 49.8%) and TDF/XTC/DTG (n = 161, 52.3%) groups. The TDF/XTC/DTG group had more participants (n = 155, 50.3%) with recent viral load at baseline < 10,000 copies/ml than the AZT/XTC/DTG (n = 80, 36.9%) and AZT/XTC/LPV/r (n = 260, 37.7%) groups. Time from most recent viral load to second-line switch was a median (IQR) of 50 days (28, 95) in the AZT/XTC/LPV/r group, 49 days (28, 102) in the AZT/XTC/DTG group and 34 days (0, 79) in the TDF/XTC/DTG group. A higher proportion of participants in the AZT/XTC/LPV/r (n = 264, 38.3%) and AZT/XTC/DTG (n = 94, 43.3%) groups had the most recent CD4 count ≤ 200 cells/μL, compared to the TDF/XTC/DTG group (n = 79, 25.6%).

**Table 1.** Baseline characteristics of PLHIV who were switched to second-line ART after virologic failure while receiving EFV^a^ or NVP-based^a^ first-line treatment

|  |  |  |  |  |
| --- | --- | --- | --- | --- |
|  | Second-line ART regimen combination |  |  |  |
| Variable | Overall,<br>N = 1214 | AZT/XTC/LPV/r,<br>N = 689 | AZT/XTC/DTG,<br>N = 217 | TDF/XTC/DTG,<br>N = 308 |
| Age in years, median (IQR) | 36 (30, 42) | 35 (30, 41) | 37 (32, 43) | 36 (30, 43) |
| Age in years |  |  |  |  |
| 15-24 | 91 (7.5%) | 54 (7.8%) | 10 (4.6%) | 27 (8.8%) |
| 25-34 | 429 (35.3%) | 255 (37.0%) | 71 (32.7%) | 103 (33.4%) |
| 35-44 | 479 (39.5%) | 274 (39.8%) | 88 (40.6%) | 117 (38.0%) |
| 45+ | 215 (17.7%) | 106 (15.4%) | 48 (22.1%) | 61 (19.8%) |
| Gender |  |  |  |  |
| Male | 485 (40.0%) | 229 (33.2%) | 109 (50.2%) | 147 (47.7%) |
| Female | 729 (60.0%) | 460 (66.8%) | 108 (49.8%) | 161 (52.3%) |
| Known pregnant (females only) | 14 (1.9%) | 10 (2.2%) | 1 (0.9%) | 3 (1.9%) |
| Known tuberculosis | 24 (2.0%) | 14 (2.0%) | 6 (2.8%) | 4 (1.3%) |
| Baseline time-period of second-line switch |  |  |  |  |
| Dec19-Feb20 | 224 (18.5%) | 190 (27.6%) | 7 (3.2%) | 27 (8.8%) |
| Mar20-May20 | 324 (26.7%) | 204 (29.6%) | 54 (24.9%) | 66 (21.4%) |
| Jun20-Aug20 | 370 (30.5%) | 165 (23.9%) | 70 (32.3%) | 135 (43.8%) |
| Sep20-Nov20 | 296 (24.4%) | 130 (18.9%) | 86 (39.6%) | 80 (26.0%) |
| Recent viral load (copies/ml) before second-line switch |  |  |  |  |
| 1,000 to <10,000 | 495 (40.8%) | 260 (37.7%) | 80 (36.9%) | 155 (50.3%) |
| 10,000 to <50,000 | 386 (31.8%) | 220 (31.9%) | 70 (32.3%) | 96 (31.2%) |
| 50000 to <100,000 | 133 (11.0%) | 80 (11.6%) | 32 (14.7%) | 21 (6.8%) |
| 100,000+ | 200 (16.5%) | 129 (18.7%) | 35 (16.1%) | 36 (11.7%) |
| Days since recent viral load (copies/ml) before second-line switch, median (IQR) | 47 (26, 92) | 50 (28, 95) | 49 (28, 102) | 34 (0, 79) |
| Days since first high viral load (copies/ml) before second-line switch, median (IQR) | 195 (140, 276) | 196 (139, 282) | 198 (141, 300) | 190 (140, 252) |
| Recent CD4 count (cells/μl) |  |  |  |  |
| ≤ 200 | 437 (36.0%) | 264 (38.3%) | 94 (43.3%) | 79 (25.6%) |
| 201–350 | 307 (25.3%) | 163 (23.7%) | 51 (23.5%) | 93 (30.2%) |
| 351–500 | 174 (14.3%) | 90 (13.1%) | 25 (11.5%) | 59 (19.2%) |
| > 500 | 133 (11.0%) | 72 (10.4%) | 19 (8.8%) | 42 (13.6%) |
| Missing | 163 (13.4%) | 100 (14.5%) | 28 (12.9%) | 35 (11.4%) |
| Days since recent CD4 count (cells/μl), median (IQR) | 400 (105, 923) | 402 (104, 928) | 273 (54, 914) | 434 (168, 914) |
| Previous first-line ART before second-line switch |  |  |  |  |
| TDF/XTC/EFV | 1,198 (98.7%) | 681 (98.8%) | 215 (99.1%) | 302 (98.1%) |
| TDF/XTC/NVP | 16 (1.3%) | 8 (1.2%) | 2 (0.9%) | 6 (1.9%) |
| ART pick-up point at baseline |  |  |  |  |
| Main clinic | 1,192 (98.2%) | 681 (98.8%) | 215 (99.1%) | 296 (96.1%) |
| CCMDD<sup>b</sup> | 22 (1.8%) | 8 (1.2%) | 2 (0.9%) | 12 (3.9%) |
| Years since ART initiation, median (IQR) | 2.9 (1.5, 5.5) | 2.9 (1.5, 5.5) | 3.5 (1.5, 6.2) | 2.6 (1.4, 4.7) |
| Years since ART initiation |  |  |  |  |
| < 2 year | 446 (36.7%) | 252 (36.6%) | 72 (33.2%) | 122 (39.6%) |
| ≥ 2 years | 768 (63.3%) | 437 (63.4%) | 145 (66.8%) | 186 (60.4%) |
| Data are n (%) or median (IQR). All percentages were calculated with the total number in the respective column headers as the denominators except otherwise stated. <sup>a</sup>Efavirenz or nevirapine based first-line regimens were in combination with TDF plus XTC. <sup>b</sup>CCMDD included external or internal pickup points, spaced fast lanes and adherence clubs. ART = Antiretroviral treatment, AZT = Zidovudine, DTG = Dolutegravir, EFV = Efavirenz, CCMDD = Central Chronic Medicines Dispensing and Distribution, IQR = Interquartile range, LVP/r = Lopinavir-ritonavir, μl = Microliter, ml = Milliliter, NVP = Nevirapine, PLHIV = People living with HIV, TDF = Tenofovir disoproxil fumarate, XTC = Emtricitabine or Lamivudine. |  |  |  |  |

During follow-up, 10.0% (n = 121) changed their originally prescribed second-line regimen after a median of 158 days, IQR (84, 234) (Table 2). By 12 months, 941 (77.5%) were retained-in-care, 80 (6.6%) had transferred out to another clinic, 16 (1.3%) were known to have died, and 177 (14.6%) were lost to follow-up. Retention-in-care at 12 months was 75.2% (n = 518) in participants receiving AZT/XTC/LPV/r, 85.7% (n = 186) in those receiving AZT/XTC/DTG and 76.9% (n = 237) in those receiving TDF/XTC/DTG (Table 3). After adjusting for potential confounders, retention-in-care at 12 months was more likely in participants receiving AZT/XTC/DTG (adjusted risk ratio [aRR] 1.14, 95% CI 1.03 to 1.27, p = 0.012; adjusted risk difference [aRD] 10.89%, 95% CI 2.01 to 19.78, p = 0.016) than those receiving AZT/XTC/LPV/r. Retention-in-care at 12 months was not different in participants receiving TDF/XTC/DTG (aRR 1.01, 95% CI 0.94 to 1.10, p = 0.733; aRD 1.04%, 95% CI -5.03 to 7.12, p = 0.736) compared to those receiving AZT/XTC/LPV/r. Retention-in-care at 12 months was lower in participants receiving TDF/XCT/DTG (76.9%) than AZT/XTC/DTG (85.7%), but the difference was not statistically significant (aRR 0.89, 95% CI 0.78 to 1.01, p = 0.060; aRD - 9.85%, 95% CI -20.33 to 0.63, p = 0.066).

**Table 2.** Follow-up outcomes in PLHIV who were switched to second-line ART after virologic failure while receiving EFV^a^ or NVP-based^a^ first-line treatment

|  |  |  |  |  |
| --- | --- | --- | --- | --- |
|  |  | **Second-line ART regimen combination** |  |  |
| **Variable** | **Overall,**<br>**N = 1214** | **AZT/XTC/LPV/r,**<br>**N = 689** | **AZT/XTC/DTG,**<br>**N = 217** | **TDF/XTC/DTG,**<br>**N = 308** |
| **Second-line regimen-change within 12 months** | 121 (10.0%) | 59 (8.6%) | 21 (9.7%) | 41 (13.3%) |
| **Days to second-line regimen-change within 12 months, median (IQR)** | 158 (84, 234) | 146 (74, 204) | 182 (97, 231) | 160 (84, 253) |
| **Second-line regimen changed to (of participants who changed regimen within 12 months)** |  |  |  |  |
| AZT/XTC/LPV/r | 17 (14.0%) | 0 (0.0%) | 4 (19.0%) | 13 (31.7%) |
| AZT/XTC/DTG | 35 (28.9%) | 16 (27.1%) | 0 (0.0%) | 19 (46.3%) |
| TDF/XTC/DTG | 26 (21.5%) | 12 (20.3%) | 14 (66.7%) | 0 (0.0%) |
| Other | 43 (35.5%) | 31 (52.5%) | 3 (14.3%) | 9 (22.0%) |
| **Follow-up outcome at 12 months** |  |  |  |  |
| Lost to follow-up | 177 (14.6%) | 112 (16.3%) | 20 (9.2%) | 45 (14.6%) |
| Died | 16 (1.3%) | 9 (1.3%) | 4 (1.8%) | 3 (1.0%) |
| Transferred out to another clinic | 80 (6.6%) | 50 (7.3%) | 7 (3.2%) | 23 (7.5%) |
| Retained in care | 941 (77.5%) | 518 (75.2%) | 186 (85.7%) | 237 (76.9%) |
| **Viral load done at 12 months (of participants retained in care at 12 months)** | 799 (84.9%) | 448 (86.5%) | 150 (80.6%) | 201 (84.8%) |
| **Days to viral load (copies/ml) at 12 months (of participants retained in care at 12 months), median (IQR)** | 357 (293, 418) | 362 (299, 419) | 342 (277, 394) | 357 (296, 426) |
| **Viral load (copies/ml) at 12 months (of participants retained in care at 12 months)** |  |  |  |  |
| <50 | 420 (52.6%) | 209 (46.7%) | 89 (59.3%) | 122 (60.7%) |
| 50-199 | 102 (12.8%) | 61 (13.6%) | 20 (13.3%) | 21 (10.4%) |
| 200-999 | 75 (9.4%) | 41 (9.2%) | 20 (13.3%) | 14 (7.0%) |
| 1000+ | 202 (25.3%) | 137 (30.6%) | 21 (14.0%) | 44 (21.9%) |
| Data are n (%) or median (IQR). All percentages were calculated with the total number in the respective column headers as the denominators except otherwise stated. <sup>a</sup>Efavirenz or nevirapine based first-line regimens were in combination with TDF plus XTC. ART = Antiretroviral treatment, AZT = Zidovudine, DTG = Dolutegravir, EFV = Efavirenz, IQR = Interquartile range, LPV/r = Lopinavir-ritonavir, µl = Microliter, ml = Milliliter, NVP = Nevirapine, PLHIV = People living with HIV, TDF = Tenofovir disoproxil fumarate, XTC = Emtricitabine or Lamivudine. |  |  |  |  |

**Table 3.** Univariable and multivariable Poisson regression models of factors associated with retention-in-care at 12 months in PLHIV who were switched to second-line ART after virologic failure while receiving EFV^a^ or NVP-based^a^ first-line treatment (N = 1214)

|  |  |  |  |  |  |  |
| --- | --- | --- | --- | --- | --- | --- |
| Variable | Level | Retention-in-care at 12 months<br>n/N (%) | Unadjusted RR<br>(95% CI) | P value | Adjusted RR<sup>b</sup><br>(95% CI) | P value |
| Second-line regimen | AZT/XTC/LPV/r | 518/689 (75.2) | 1 | - | 1 | - |
|  | AZT/XTC/DTG | 186/217 (85.7) | 1.14 (1.03-1.27) | 0.013 | 1.14 (1.03-1.27) | 0.012 |
|  | TDF/XTC/DTG | 237/308 (76.9) | 1.02 (0.94-1.11) | 0.630 | 1.01 (0.94-1.10) | 0.733 |
| Age at baseline | 15-24 | 67/91 (73.6) | 1 | - | 1 | - |
|  | 25-34 | 323/429 (75.3) | 1.03 (0.90-1.17) | 0.714 | 1.03 (0.90-1.17) | 0.714 |
|  | 35-44 | 377/479 (78.7) | 1.07 (0.94-1.22) | 0.307 | 1.07 (0.95-1.22) | 0.274 |
|  | 45+ | 174/215 (80.9) | 1.10 (0.98-1.25) | 0.113 | 1.10 (0.97-1.24) | 0.135 |
| Gender | Male | 373/485 (76.9) | 1 | - | 1 | - |
|  | Female | 568/729 (77.9) | 1.01 (0.95-1.08) | 0.673 | 1.03 (0.98-1.10) | 0.258 |
| Known tuberculosis status at baseline | No | 925/1190 (77.7) | 1 | - | 1 | - |
|  | Yes | 16/24 (66.7) | 0.85 (0.63-1.15) | 0.299 | 0.86 (0.64-1.15) | 0.312 |
| Recent viral load (copies/ml) at baseline | 1000 to < 10000 | 399/495 (80.6) | 1 | - | 1 | - |
|  | ≥ 10000 | 542/719 (75.4) | 0.93 (0.88-0.99) | 0.030 | 0.94 (0.88-1.00) | 0.042 |
| Recent CD4 count (cells/μl) at baseline | ≤ 200 | 338/437 (77.3) | 1 | - | - | - |
|  | 201–350 | 235/307 (76.5) | 0.99 (0.91-1.08) | 0.832 | - | - |
|  | 351–500 | 128/174 (73.6) | 0.95 (0.86-1.05) | 0.327 | - | - |
|  | > 500 | 106/133 (79.7) | 1.03 (0.92-1.15) | 0.585 | - | - |
|  | Missing | 134/163 (82.2) | 1.08 (0.99-1.17) | 0.067 | - | - |
| Years on ART at baseline | < 2 year | 335/446 (75.1) | 1 | - | - | - |
|  | ≥ 2 years | 606/768 (78.9) | 1.05 (0.97-1.13) | 0.207 | - | - |
| Data are n/N (%), unless otherwise stated. <sup>a</sup>Efavirenz or nevirapine based first-line regimens were in combination with TDF plus XTC. <sup>b</sup>The primary exposure effect (retention-in-care at 12 months) is adjusted for all other variables in the table as potential confounders except CD4 count and Years on ART at baseline. ART = Antiretroviral treatment, AZT = Zidovudine, DTG = Dolutegravir, EFV = Efavirenz, LVP/r = Lopinavir-ritonavir, μl = Microliter, ml = Milliliter, NVP = Nevirapine, PLHIV = People living with HIV, RR = Risk ratio, TDF = Tenofovir disoproxil fumarate, XTC = Emtricitabine or Lamivudine. |  |  |  |  |  |  |

Of 941 participants who were retained in care at 12 months, 799 (84.9%) had a viral load done at a median of 357 days, IQR (293-418) (Table 2). By regimen, 448 (86.5%) of those receiving AZT/XTC/LPV/r, 150 (80.6%) of those receiving AZT/XTC/DTG and 201 (84.8%) of those receiving TDF/XTC/DTG had a viral load done at 12 months follow-up. Of participants with a viral load at 12 months, viral suppression (< 50 copies/ml) was higher in those receiving AZT/XTC/DTG (n = 89, 59.3%) and TDF/XTC/DTG (n = 122, 60.7%) than AZT/XTC/LPV/r (n = 209, 46.7%). Viral suppression (< 50 copies/ml) at 12 months was more likely in participants receiving AZT/XTC/DTG (aRR 1.25, 95% CI 1.06 to 1.47, p = 0.009; aRD 11.57%, 95% CI 2.37 to 20.76, p = 0.014) and TDF/XTC/DTG (aRR 1.30, 95% CI 1.14 to 1.48, p < 0.001; aRD 14.16%, 95% CI 7.14 to 21.18, p < 0.001) than participants receiving AZT/XTC/LPV/r (Table 4). Viral suppression (< 50 copies/ml) at 12 months was similar in participants receiving TDF/XCT/DTG compared to AZT/XTC/DTG (aRR 1.04, 95% CI 0.88 to 1.24, p = 0.624; aRD 2.59%, 95% CI -7.78 to 12.60, p = 0.624). In a post-hoc sensitivity analysis presented as part of the supplementary results, viral suppression (< 1000 copies/ml) at 12 months was more likely in participants receiving AZT/XTC/DTG (86.0%, aRR 1.19, 95% CI 1.07 to 1.32, p = 0.001; aRD 13.22%, 95% CI 5.02 to 21.41, p = 0.002) and TDF/XTC/DTG (78.1%, aRR 1.11, 95% CI 1.01 to 1.22, p = 0.033; aRD 7.63%, 95% CI 0.50 to 14.77, p = 0.036) than participants receiving AZT/XTC/LPV/r (69.4%) (Table S 1). Viral suppression (< 1000 copies/ml) at 12 months was similar in participants receiving TDF/XCT/DTG compared to AZT/XTC/DTG (aRR 0.93, 95% CI 0.85 to 1.02, p = 0.143; aRD -5.58%, 95% CI -13.12 to 1.95, p = 0.146).

**Table 4.** Univariable and multivariable Poisson regression models of factors associated with viral suppression (< 50 copies/ml) at 12 months in PLHIV who were switched to second-line ART after virologic failure while receiving EFV^a^ or NVP-based^a^ first-line treatment (N = 799)

|  |  |  |  |  |  |  |
| --- | --- | --- | --- | --- | --- | --- |
| Variable | Level | Viral load at 12 months<br>< 50 copies/ml<br>n/N (%) | Unadjusted RR<br>(95% CI) | P value | Adjusted RR<sup>b</sup><br>(95% CI) | P value |
| Second-line regimen | AZT/XTC/LPV/r | 209/448 (46.7) | 1 | - | 1 | - |
|  | AZT/XTC/DTG | 89/150 (59.3) | 1.22 (1.03-1.46) | 0.022 | 1.25 (1.06-1.47) | 0.009 |
|  | TDF/XTC/DTG | 122/201 (60.7) | 1.31 (1.15-1.49) | <0.001 | 1.30 (1.14-1.48) | <0.001 |
| Age at baseline | 15-24 | 21/56 (37.5) | 1 | - | 1 | - |
|  | 25-34 | 153/282 (54.3) | 1.46 (0.99-2.14) | 0.056 | 1.50 (1.01-2.21) | 0.043 |
|  | 35-44 | 159/308 (51.6) | 1.37 (0.91-2.06) | 0.127 | 1.45 (0.96-2.17) | 0.075 |
|  | 45+ | 87/153 (56.9) | 1.55 (1.06-2.27) | 0.024 | 1.58 (1.07-2.33) | 0.022 |
| Gender | Male | 156/313 (49.8) | 1 | - | 1 | - |
|  | Female | 264/486 (54.3) | 1.10 (0.98-1.23) | 0.111 | 1.12 (1.00-1.25) | 0.053 |
| Known tuberculosis status at baseline | No | 415/784 (52.9) | 1 | - | 1 | - |
|  | Yes | 5/15 (33.3) | 0.66 (0.30-1.45) | 0.302 | 0.69 (0.32-1.48) | 0.337 |
| Recent viral load (copies/ml) at baseline | 1000 to < 10000 | 198/337 (58.8) | 1 | - | 1 | - |
|  | ≥ 10000 | 222/462 (48.1) | 0.83 (0.73-0.93) | 0.002 | 0.85 (0.76-0.96) | 0.008 |
| Recent CD4 count (cells/μl) at baseline | ≤ 200 | 153/292 (52.4) | 1 | - | - | - |
|  | 201–350 | 96/194 (49.5) | 0.95 (0.81-1.11) | 0.492 | - | - |
|  | 351–500 | 66/108 (61.1) | 1.14 (0.94-1.39) | 0.180 | - | - |
|  | > 500 | 42/90 (46.7) | 0.90 (0.66-1.23) | 0.514 | - | - |
|  | Missing | 63/115 (54.8) | 1.04 (0.83-1.31) | 0.730 | - | - |
| Years on ART at baseline | < 2 year | 156/286 (54.5) | 1 | - | - | - |
|  | ≥ 2 years | 264/513 (51.5) | 0.95 (0.82-1.10) | 0.479 | - | - |
| Data are n/N (%), unless otherwise stated. <sup>a</sup>Efavirenz or nevirapine based first-line regimens were in combination with TDF plus XTC. <sup>b</sup>The primary exposure effect (viral suppression at 12 months) is adjusted for all other variables in the table as potential confounders. ART = Antiretroviral treatment, AZT = Zidovudine, DTG = Dolutegravir, EFV = Efavirenz, LPV/r = Lopinavir-ritonavir, μl = Microliter, ml = Milliliter, NVP = Nevirapine, PLHIV = People living with HIV, RR = Risk ratio, TDF = Tenofovir disoproxil fumarate, XTC = Emtricitabine or Lamivudine. |  |  |  |  |  |  |

The supplementary results (Tables S2, S3, and S4) showed no significant confounding of retention-in-care and viral suppression outcomes by recent baseline CD4 count and time on ART at baseline. In Tables S5, S6 and S7, results show that retention-in-care and viral suppression outcomes were consistent with the main analysis after excluding participants who changed their originally prescribed second-line regimen within 12 months after baseline.

## DISCUSSION

In this retrospective cohort study with routine data from 59 clinics in South Africa, compared to second-line LPV/r-based regimens, second-line DTG-based regimens were associated with similar or better retention in care and better viral suppression. We did not find evidence of a significant difference in retention or viral suppression between TDF/XTC/DTG and AZT/XTC/DTG.

We evaluated retention-in-care at 12 months because drug tolerability is known to influence adherence^15^ and retention-in-care^16^. We saw higher retention-in-care with AZT/XTC/DTG than AZT/XTC/LPV/r consistent with the favourable safety profile of DTG-based versus LPV/r-based regimens for second-line treatment in the DAWNING trial^5^ and generally reported during first-line treatment^5, 17–19^. Retention-in-care with TDF/XTC/DTG (76.9%) was lower than with AZT/XTC/DTG (85.7%) although not significantly different (P value = 0.066), but we expected similar rates as TDF is slightly more tolerable than AZT^20, 21^. The week-96 results of the NADIA trial^22^ showed identical low rates of adverse events leading to second-line treatment discontinuation in the TDF-based (n = 2, 1.0%) and the AZT-based (n = 3, 1.0%) groups.

The DAWNING trial is the only clinical trial directly comparing the efficacy of DTG versus LPV/r for second-line ART. The trial enrolled 624 PLHIV ≥ 18 years with virologic failure during first-line treatment and randomized 312 to receive DTG and 312 to receive LPV/r in a second-line regimen plus two NRTIs, with at least one being fully active.^5^ Most participants in the DTG-based group reported high ART adherence scores and lower treatment-related adverse events (67.0% and 16.0%) compared to the LPV/r group (56.0% and 38.0%).^5^ There were also fewer adverse events leading to treatment discontinuation in the DTG group (3.0%) than the LPV/r group (6.0%), which may explain the improved retention-in-care that we found with AZT/XTC/DTG versus AZT/XTC/LPV/r.^5^ In the primary intention to treat analysis, the primary outcome of viral suppression (viral load < 50 copies/ml) at 48 weeks was higher in the DTG group (84.0%) compared to the LPV/r group (70.0%), adjusted difference 13.8%; 95% CI 7.3 to 20.3.^5^

There are four clinical trials assessing the efficacy of recycling TDF in a second-line regimen. The NADIA trial used a 2 x 2 factorial design to randomise PLHIV with virologic failure during first-line treatment to receive second-line dolutegravir or lopinavir-boosted darunavir and either tenofovir or zidovudine.^6^ Recycling tenofovir for second-line treatment was non-inferior to switching to zidovudine for viral suppression (viral load < 400 copies/ml) at 48 weeks.^6^ Consistent with results from the NADIA trial, we found no difference between TDF/XTC/DTG versus AZT/XTC/DTG for viral suppression at < 50 copies/ml. The smaller single-arm ARTIST trial in 62 participants showed 74.0% viral suppression (< 50 copies/ml) at 48 weeks with TDF/XTC/DTG during second-line treatment.^23, 24^ Preliminary results from the VISEND^25^ and D2EFT^26^ trials also found TDF/XTC/DTG as non-inferior to ritonavir-boosted lopinavir or atazanavir (VISEND) and darunavir (D2FT). In this routine care setting, TDF/XTC/DTG was associated with better viral suppression versus AZT/XTC/LPV/r.

Viral suppression rates are generally higher in these trials than we found in routine care, probably due to better treatment adherence and monitoring^27, 28^ among participants in clinical trials^29^. But differences in cohort baseline virologic failure and post-baseline viral suppression thresholds might also be responsible for the different outcomes. Although the DAWNING^5^ trial used a viral suppression of < 50 copies/ml, it included participants with a baseline viral load between 400 to < 1000 copies/ml (9.0% in the DTG group, 11.0% in the LPV/r group) versus our cohort which used a guideline-defined threshold of ≥ 1000 copies/ml. The NADIA^6^ trial used a baseline viral load of ≥ 1000 copies/ml as we did but defined viral suppression at < 400 copies/ml. The VISEND^25^ trial included participants with a baseline viral load of ≥ 1000 copies/ml and used a viral suppression threshold of < 1000 copies/ml. The resulting viral suppression < 1000 copies/ml at 12 months (82.0% with TDF/XTC/DTG and 76.0% with AZT/3TC plus LPV/r or atazanavir/r)^25^ was similar to what we found in post-hoc sensitivity analyses with same thresholds (78.1% with TDF/XTC/DTG, 69.4% with AZT/XTC/LPV/r and 86.0% with AZT/XTC/DTG). A multisite cohort study conducted between 2007 to 2009 in 6 African countries, including South Africa, reported 13.9% virologic failure (≥ 400 copies/ml) at 12 months after starting second-line treatment with protease-inhibitor-based regimens, which we interpret as 86.1% viral suppression (< 400 copies/ml).^27^

Overall, outcomes were poor in this cohort of people switching to second-line ART after first-line virologic failure in routine care. Of the 1214 people, just about a third, 420 (34.6%), achieved programmatic retention-in-care and viral suppression milestones at 12 months. This highlights the need to improve other outcomes in the care cascade in ART programmes during second-line treatment, particularly adherence counselling, as regimen choice is only one factor necessary for improving HIV treatment outcomes.

Our findings are among the first evidence of outcomes with two common dolutegravir-based regimen combinations for second-line ART in resource-limited routine healthcare settings. We used guideline-defined virologic failure, viral suppression, and retention-in-care and adjusted for the effects of baseline characteristics when switching to second-line treatment. Our findings support WHO’s recommendation of dolutegravir for second-line ART in adults with treatment failure on a first-line regimen containing an NNRTI such as nevirapine or efavirenz.^1^

Furthermore, WHO recommends the substitution of TDF, a common drug in most first-line regimens in LMICs, with zidovudine when switching to second-line treatment to ensure having an active NRTI backbone due to limited resistance testing^30^ for selecting appropriate NRTIs.^1, 2^ However, based on results from the NADIA trial suggesting the non-inferiority of recycling TDF instead of switching to AZT, and the availability of TLD as a fixed dose combination, TDF/XTC/DTG is considered an easily implementable regimen.^7^ Our findings have provided further assurance regarding these assertions with evidence from routine care that recycled TDF in a second-line DTG-based regimen can result in similar viral suppression <50 copies/ml at 12 months as with switching to AZT, both of which can be more effective than the previous regimen of AZT/XTC/LPV/r. This finding is, therefore, also relevant to other resource-limited settings where resistance testing is not routinely done to guide the selection of NRTIs for second-line treatment.

Our analysis had some potential limitations. First, we used data from only one district in South Africa, which might have limited the generalizability of the findings, however, the sample size was large considering the high HIV burden in our setting. Second, we only assessed 12-month outcomes, and evaluating longer-term follow-up will be important in future analyses. Third, although we adjusted for the most relevant baseline characteristics, we cannot rule out potential unmeasured confounders. Fourth, we were unable to include the recent CD4 count and time on ART in the multivariable analysis as it led to overfitted models with predicted probabilities exceeding one. We, therefore, evaluated the impact of baseline CD4 count and years on ART in supplementary analyses, which showed no evidence of significant confounding of the primary outcomes. Fifth, in a new era of DTG, clinicians and nurses might have selected specific PLHIV for DTG treatment who were more likely to have better outcomes. Furthermore, people who received TDF/XTC/DTG after virological failure may have been put on this regimen in error as part of the transition to first-line dolutegravir or were more likely to be anaemic, a contraindication to AZT^4^. They may, therefore, not have received similar treatment to those receiving the recommended second-line regimens (for example, they may not have received enhanced adherence counselling), which could make them different from the AZT groups introducing further bias. Furthermore, we do not have follow-up measures of regimen-related adverse events for comparison, but the DAWNING trial^5^ showed a favourable safety profile with DTG than LPV/r during second-line ART, and the NADIA trial showed that recycled TDF and switching to AZT for second-line treatment are both safe^22^.

In conclusion, we found that among people who experienced virologic failure during first-line non-dolutegravir-based ART, dolutegravir use for second-line treatment resulted in similar or better retention-in-care and better viral suppression at 12 months follow-up than the previous ritonavir-boosted lopinavir regimen all used in combination with AZT plus XTC. Furthermore, recycled TDF plus XTC with dolutegravir for second-line treatment yielded identical retention-in-care and viral suppression impact as with AZT plus XTC. These findings support the ongoing use of DTG-based second-line regimens in low- and middle-income countries.

## Supporting information

Supplementary results

## Data Availability

We cannot publicly share the data used for this analysis because of the legal and ethical requirements regarding the use of routinely collected clinical data in South Africa. Interested parties can request access to the data from the eThekwini Municipality Health Unit and the South African National Department of Health TB/HIV Information System (contact details obtainable upon request to JD).

## ABBREVIATIONS

ART: Anti-retroviral therapy
RD: Risk difference
RR: Risk Ratio
LMIC: Low- and Middle-Income Country
PLHIV: People living with HIV
WHO: World Health Organization

## Contributors

KA, YS, LL, RJL, KN, NG and JD conceptualised the study. TK, YS, and RvH oversaw data collection. TK, and JvdM managed data curation. KA, YS, TK, JvdM, LL, RvH, NG and JD had full access to the data in the study through their role in eThekwini Municipality, the Health Informatics Directorate, or permissions granted to the Centre for the AIDS Programme of Research in South Africa. KA, JvdM, LL, and JD analysed the data. KA drafted the manuscript. All authors contributed to the interpretation of results, critically reviewed, and approved the final version for submission.

## Acknowledgements

We thank the eThekwini Municipality Health Unit staff, patients, and primary care clinics. This work was supported by funding from the Africa Oxford Initiative (AfiOx-160), the International Association of Providers of AIDS Care (2021-ISG-Y1-10004), and the Bill & Melinda Gates Foundation (INV-051067). Under the grant conditions of the Bill & Melinda Gates Foundation, a Creative Commons Attribution 4.0 Generic License has already been assigned to the Author Accepted Manuscript version that might arise from this submission. JD, Academic Clinical Lecturer is funded by the NIHR for this research project. The views expressed in this publication are those of the author (s) and not necessarily those of the NIHR, NHS or the UK Department of Health and Social Care.

