## Supplementary results for "Clinical outcomes after the introduction of dolutegravir for second-line antiretroviral therapy in South Africa: a retrospective cohort study"

Appendix A: Supplementary Tables

Table S 1. Post-hoc sensitivity analysis: Univariable and multivariable Poisson regression models of factors associated with viral suppression (< 1000 copies/ml) at 12 months (N = 799)

Table S 2. Potential confounding effects of CD4 count and Years on ART on the risk ratios of 12-month retention-in-care (N = 1214)

Table S 3. Potential confounding effects of CD4 count and Years on ART on the risk ratios of 12-month viral suppression (< 50 copies/ml) (N = 799)

Table S 4. Potential confounding effects of CD4 count and Years on ART on the risk ratios of 12-month viral suppression (< 1000 copies/ml) (N = 799)

Table S 5. Retention-in-care at 12 months excluding 121 participants who changed their originally prescribed second-line regimen within 12 months after baseline (N = 1093)

Table S 6. Viral suppression (<50 copies/ml) at 12 months excluding 121 participants who changed their originally prescribed second-line regimen within 12 months after baseline (N = 713)

Table S 7. Viral suppression (<1000 copies/ml) at 12 months excluding 121 participants who changed their originally prescribed second-line regimen within 12 months after baseline (N = 713)

| **Table S 1. Post-hoc sensitivity analysis: Univariable and multivariable Poisson regression models of factors associated with viral suppression (< 1000 copies/ml) at 12 months (N = 799)** | | | | | | |
|---|---|---|---|---|---|---|
| Variable | Level | Viral load at 12 months < 1000 copies/ml n/N (%) | Unadjusted RR (95% CI) | P value | Adjusted RR[b] (95% CI) | P value |
| Second-line regimen | AZT/XTC/LPV/r | 311/448 (69.4) | 1 | - | 1 | - |
| | AZT/XTC/DTG | 129/150 (86.0) | 1.18 (1.07-1.31) | 0.001 | 1.19 (1.07-1.32) | 0.001 |
| | TDF/XTC/DTG | 157/201 (78.1) | 1.13 (1.02-1.24) | 0.015 | 1.11 (1.01-1.22) | 0.033 |
| Age at baseline | 15-24 | 31/56 (55.4) | 1 | - | 1 | - |
| | 25-34 | 207/282 (73.4) | 1.32 (1.02-1.70) | 0.035 | 1.35 (1.04-1.74) | 0.024 |
| | 35-44 | 240/308 (77.9) | 1.38 (1.04-1.82) | 0.024 | 1.40 (1.06-1.84) | 0.019 |
| | 45+ | 119/153 (77.8) | 1.42 (1.09-1.85) | 0.010 | 1.37 (1.05-1.81) | 0.022 |
| Gender | Male | 243/313 (77.6) | 1 | - | 1 | - |
| | Female | 354/486 (72.8) | 0.95 (0.88-1.02) | 0.14 | 0.95 (0.89-1.03) | 0.22 |
| Known tuberculosis status at baseline | No | 588/784 (75.0) | 1 | - | 1 | - |
| | Yes | 9/15 (60.0) | 0.82 (0.54-1.25) | 0.36 | 0.81 (0.55-1.22) | 0.32 |
| Recent viral load (copies/ml) at baseline | 1000 to < 10000 | 276/337 (81.9) | 1 | - | 1 | - |
| | ≥ 10000 | 321/462 (69.5) | 0.86 (0.79-0.93) | <0.001 | 0.85 (0.78-0.92) | <0.001 |
| Recent CD4 count (cells/µL) at baseline | ≤ 200 | 220/292 (75.3) | 1 | - | 1 | - |
| | 201–350 | 142/194 (73.2) | 0.98 (0.90-1.06) | 0.58 | - | - |
| | 351–500 | 88/108 (81.5) | 1.07 (0.94-1.22) | 0.30 | - | - |
| | > 500 | 61/90 (67.8) | 0.91 (0.76-1.09) | 0.31 | - | - |
| | Missing | 86/115 (74.8) | 1.00 (0.88-1.14) | 0.99 | - | - |
| Years on ART at baseline | < 2 year | 220/286 (76.9) | - | - | - | - |
| | ≥ 2 years | 377/513 (73.5) | 0.96 (0.89-1.03) | 0.27 | - | - |

Data are n/N (%), unless otherwise stated. [a]Efavirenz or nevirapine based first-line regimens were in combination with TDF plus XTC. [b]The primary exposure effect (viral suppression at 12 months) is adjusted for all other variables in the table as potential confounders. ART = Antiretroviral treatment, AZT = Zidovudine, DTG = Dolutegravir, EFV = Efavirenz, LVP/r = Lopinavir-ritonavir, µl = Microliter, ml = Milliliter, NVP = Nevirapine, PLHIV = People living with HIV, RR = Risk ratio, TDF = Tenofovir disoproxil fumarate, XTC = Emtricitabine or Lamivudine.

| Table S 2. Potential confounding effects of CD4 count and Years on ART on the risk ratios of 12-month retention-in-care (N = 1214) | | | | | | |
|---|---|---|---|---|---|---|
| Variable | Level | Retention-in-care at 12 months n/N (%) | Unadjusted RR (95% CI) | P value | Adjusted RR[a] (95% CI) | P value |
| Second-line regimen | AZT/XTC/LPV/r | 518/689 (75.2) | 1 | - | 1 | - |
| | AZT/XTC/DTG | 186/217 (85.7) | 1.14 (1.03-1.27) | 0.013 | 1.14 (1.02-1.27) | 0.016 |
| | TDF/XTC/DTG | 237/308 (76.9) | 1.02 (0.94-1.11) | 0.63 | 1.03 (0.95-1.12) | 0.52 |
| Recent CD4 count (cells/μL) at baseline | ≤ 200 | 338/437 (77.3) | 1 | - | 1 | - |
| | 201–350 | 235/307 (76.5) | 0.99 (0.91-1.08) | 0.83 | 0.99 (0.90-1.08) | 0.76 |
| | 351–500 | 128/174 (73.6) | 0.95 (0.86-1.05) | 0.33 | 0.95 (0.86-1.04) | 0.28 |
| | > 500 | 106/133 (79.7) | 1.03 (0.92-1.15) | 0.59 | 1.03 (0.92-1.15) | 0.63 |
| | Missing | 134/163 (82.2) | 1.08 (0.99-1.17) | 0.067 | 1.08 (1.00-1.17) | 0.05 |
| Years on ART at baseline | < 2 year | 335/446 (75.1) | 1 | - | 1 | - |
| | ≥ 2 years | 606/768 (78.9) | 1.05 (0.97-1.13) | 0.21 | 1.05 (0.97-1.14) | 0.20 |

Data are n/N (%), unless otherwise stated. [a]The primary exposure effect (retention-in-care at 12 months) is adjusted for CD4 count and Years on ART at baseline. ART = Antiretroviral treatment, AZT = Zidovudine, DTG = Dolutegravir, LVP/r = Lopinavir-ritonavir, TDF = Tenofovir disoproxil fumarate, RR = Risk ratio, XTC = Emtricitabine or Lamivudine.

**Table S 3. Potential confounding effects of CD4 count and Years on ART on the risk ratios of 12-month viral suppression (< 50 copies/ml) (N = 799)**

| Variable | Level | Viral load at 12 months < 50 copies/ml n/N (%) | Unadjusted RR (95% CI) | P value | Adjusted RR[a] (95% CI) | P value |
|---|---|---|---|---|---|---|
| Second-line regimen | AZT/XTC/LPV/r | 209/448 (46.7) | 1 | - | 1 | - |
| | AZT/XTC/DTG | 89/150 (59.3) | 1.22 (1.03-1.46) | 0.022 | 1.23 (1.04-1.47) | 0.017 |
| | TDF/XTC/DTG | 122/201 (60.7) | 1.31 (1.15-1.49) | <0.001 | 1.31 (1.15-1.49) | <0.001 |
| Recent CD4 count (cells/µL) at baseline | ≤ 200 | 153/292 (52.4) | 1 | - | 1 | - |
| | 201–350 | 96/194 (49.5) | 0.95 (0.81-1.11) | 0.49 | 0.94 (0.80-1.10) | 0.43 |
| | 351–500 | 66/108 (61.1) | 1.14 (0.94-1.39) | 0.18 | 1.13 (0.92-1.38) | 0.23 |
| | > 500 | 42/90 (46.7) | 0.90 (0.66-1.23) | 0.51 | 0.88 (0.64-1.20) | 0.42 |
| | Missing | 63/115 (54.8) | 1.04 (0.83-1.31) | 0.73 | 1.05 (0.84-1.31) | 0.69 |
| Years on ART at baseline | < 2 year | 156/286 (54.5) | 1 | - | 1 | - |
| | ≥ 2 years | 264/513 (51.5) | 0.95 (0.82-1.10) | 0.48 | 0.94 (0.81-1.10) | 0.47 |

Data are n/N (%), unless otherwise stated. [a]The primary exposure effect (viral suppression at 12 months) is adjusted for CD4 count and Years on ART at baseline. ART = Antiretroviral treatment, AZT = Zidovudine, DTG = Dolutegravir, LVP/r = Lopinavir-ritonavir, TDF = Tenofovir disoproxil fumarate, RR = Risk ratio, XTC = Emtricitabine or Lamivudine.

**Table S 4. Potential confounding effects of CD4 count and Years on ART on the risk ratios of 12-month viral suppression (< 1000 copies/ml) (N = 799)**

| Variable | Level | Viral load at 12 months < 1000 copies/ml n/N (%) | Unadjusted RR (95% CI) | P value | Adjusted RR[a] (95% CI) | P value |
|---|---|---|---|---|---|---|
| Second-line regimen | AZT/XTC/LPV/r | 311/448 (69.4) | 1 | - | 1 | - |
| | AZT/XTC/DTG | 129/150 (86.0) | 1.18 (1.07-1.31) | 0.001 | 1.19 (1.07-1.32) | 0.001 |
| | TDF/XTC/DTG | 157/201 (78.1) | 1.13 (1.02-1.24) | 0.015 | 1.13 (1.02-1.25) | 0.018 |
| Recent CD4 count (cells/µL) at baseline | ≤ 200 | 220/292 (75.3) | 1 | - | 1 | - |
| | 201–350 | 142/194 (73.2) | 0.98 (0.90-1.06) | 0.58 | 0.98 (0.90-1.07) | 0.67 |
| | 351–500 | 88/108 (81.5) | 1.07 (0.94-1.22) | 0.30 | 1.08 (0.94-1.23) | 0.27 |
| | > 500 | 61/90 (67.8) | 0.91 (0.76-1.09) | 0.31 | 0.91 (0.76-1.09) | 0.30 |
| | Missing | 86/115 (74.8) | 1.00 (0.88-1.14) | 0.99 | 1.01 (0.88-1.15) | 0.93 |
| Years on ART at baseline | < 2 year | 220/286 (76.9) | 1 | - | 1 | - |
| | ≥ 2 years | 377/513 (73.5) | 0.96 (0.89-1.03) | 0.26 | 0.95 (0.88-1.03) | 0.22 |

Data are n/N (%), unless otherwise stated. [a]The primary exposure effect (viral suppression at 12 months) is adjusted for CD4 count and Years on ART at baseline. ART = Antiretroviral treatment, AZT = Zidovudine, DTG = Dolutegravir, LVP/r = Lopinavir-ritonavir, TDF = Tenofovir disoproxil fumarate, RR = Risk ratio, XTC = Emtricitabine or Lamivudine.

**Table S 5. Univariable and multivariable Poisson regression models of factors associated with retention-in-care at 12 months excluding 121 participants who changed their originally prescribed second-line regimen within 12 months after baseline (N = 1093)**

| Variable | Level | Retention-in-care at 12 months n/N (%) | Unadjusted RR (95% CI) | P value | Adjusted RR[a] (95% CI) | P value |
|---|---|---|---|---|---|---|
| Second-line regimen | AZT/XTC/LPV/r | 469/630 (74.4) | 1 | - | 1 | - |
| | AZT/XTC/DTG | 170/196 (86.7) | 1.17 (1.05-1.31) | 0.004 | 1.18 (1.06-1.31) | 0.003 |
| | TDF/XTC/DTG | 200/267 (74.9) | 1.00 (0.91-1.10) | 0.96 | 1.00 (0.91-1.09) | 0.98 |
| Age at baseline | 15-24 | 60/81 (74.1) | 1 | - | 1 | - |
| | 25-34 | 296/397 (74.6) | 1.01 (0.88-1.16) | 0.90 | 1.01 (0.88-1.16) | 0.90 |
| | 35-44 | 334/427 (78.2) | 1.06 (0.92-1.21) | 0.43 | 1.06 (0.93-1.21) | 0.36 |
| | 45+ | 149/188 (79.3) | 1.08 (0.94-1.23) | 0.28 | 1.07 (0.94-1.22) | 0.29 |
| Gender | Male | 325/432 (75.2) | 1 | - | 1 | - |
| | Female | 514/661 (77.8) | 1.03 (0.97-1.11) | 0.33 | 1.05 (0.98-1.12) | 0.13 |
| Known tuberculosis at baseline | No | 828/1075 (77.0) | 1 | - | 1 | - |
| | Yes | 11/18 (61.1) | 0.79 (0.55-1.13) | 0.19 | 0.79 (0.56-1.13) | 0.20 |
| Recent viral load (copies/ml) at baseline | 1,000 to <10,000 | 353/440 (80.2) | 1 | - | 1 | - |
| | 10,000+ | 486/653 (74.4) | 0.93 (0.87-0.99) | 0.022 | 0.93 (0.87-1.00) | 0.036 |
| Recent CD4 count (cells/µl) at baseline | ≤ 200 | 303/396 (76.5) | 1 | - | - | - |
| | 201–350 | 203/269 (75.5) | 0.99 (0.90-1.08) | 0.78 | - | - |
| | 351–500 | 115/157 (73.2) | 0.96 (0.86-1.07) | 0.43 | - | - |
| | > 500 | 100/126 (79.4) | 1.04 (0.93-1.16) | 0.51 | - | - |
| | Missing | 118/145 (81.4) | 1.08 (0.99-1.17) | 0.090 | - | - |
| Years on ART at baseline | < 2 year | 302/405 (74.6) | 1 | - | - | - |
| | ≥ 2 years | 537/688 (78.1) | 1.05 (0.97-1.13) | 0.25 | - | - |

Data are n/N (%), unless otherwise stated. [a]The primary exposure effect (retention-in-care at 12 months) is adjusted for all other variables in the table as potential confounders except CD4 count and Years on ART at baseline. ART = Antiretroviral treatment, AZT = Zidovudine, DTG = Dolutegravir, EFV = Efavirenz, LVP/r = Lopinavir-ritonavir, µl = Microliter, ml = Milliliter, NVP = Nevirapine, PLHIV = People living with HIV, RR = Risk ratio, TDF = Tenofovir disoproxil fumarate, XTC = Emtricitabine or Lamivudine.

**Table S 6. Univariable and multivariable Poisson regression models of factors associated with viral suppression (<50 copies/ml) at 12 months excluding 121 participants who changed their originally prescribed second-line regimen within 12 months after baseline (N = 713)**

| Variable | Level | Viral load at 12 months < 50 copies/ml n/N (%) | Unadjusted RR (95% CI) | P value | Adjusted RR[a] (95% CI) | P value |
|---|---|---|---|---|---|---|
| Second-line regimen | AZT/XTC/LPV/r | 188/407 (46.2) | 1 | - | 1 | - |
| | AZT/XTC/DTG | 84/137 (61.3) | 1.27 (1.04-1.56) | 0.019 | 1.29 (1.06-1.57) | 0.010 |
| | TDF/XTC/DTG | 103/169 (60.9) | 1.32 (1.14-1.53) | <0.001 | 1.31 (1.13-1.52) | <0.001 |
| Age at baseline | 15-24 | 19/51 (37.3) | 1 | - | 1 | - |
| | 25-34 | 140/257 (54.5) | 1.47 (0.98-2.22) | 0.065 | 1.50 (1.00-2.27) | 0.053 |
| | 35-44 | 142/275 (51.6) | 1.38 (0.90-2.12) | 0.14 | 1.48 (0.97-2.27) | 0.071 |
| | 45+ | 74/130 (56.9) | 1.58 (1.04-2.40) | 0.031 | 1.63 (1.07-2.49) | 0.023 |
| Gender | Male | 132/271 (48.7) | 1 | - | 1 | - |
| | Female | 243/442 (55.0) | 1.15 (1.00-1.32) | 0.042 | 1.17 (1.03-1.33) | 0.016 |
| Known tuberculosis at baseline | No | 373/703 (53.1) | 1 | - | 1 | - |
| | Yes | 2/10 (20.0) | 0.40 (0.12-1.31) | 0.13 | 0.41 (0.12-1.32) | 0.14 |
| Recent viral load (copies/ml) at baseline | 1,000 to <10,000 | 177/301 (58.8) | 1 | - | 1 | - |
| | 10,000+ | 198/412 (48.1) | 0.84 (0.74-0.94) | 0.003 | 0.87 (0.78-0.98) | 0.019 |
| Recent CD4 count (cells/µl) at baseline | ≤ 200 | 139/264 (52.7) | 1 | - | - | - |
| | 201–350 | 79/167 (47.3) | 0.91 (0.77-1.08) | 0.27 | - | - |
| | 351–500 | 60/96 (62.5) | 1.17 (0.98-1.40) | 0.09 | - | - |
| | > 500 | 41/86 (47.7) | 0.93 (0.68-1.27) | 0.64 | - | - |
| | Missing | 56/100 (56.0) | 1.07 (0.84-1.36) | 0.57 | - | - |
| Years on ART at baseline | < 2 year | 139/254 (54.7) | 1 | - | - | - |
| | ≥ 2 years | 236/459 (51.4) | 0.94 (0.80-1.10) | 0.43 | - | - |

Data are n/N (%), unless otherwise stated. [a]The primary exposure effect (viral suppression at 12 months) is adjusted for all other variables in the table as potential confounders except CD4 count and Years on ART at baseline. ART = Antiretroviral treatment, AZT = Zidovudine, DTG = Dolutegravir, EFV = Efavirenz, LVP/r = Lopinavir-ritonavir, µl = Microliter, ml = Milliliter, NVP = Nevirapine, PLHIV = People living with HIV, RR = Risk ratio, TDF = Tenofovir disoproxil fumarate, XTC = Emtricitabine or Lamivudine.

| **Table S 7. Univariable and multivariable Poisson regression models of factors associated with viral suppression (<1000 copies/ml) at 12 months excluding 121 participants who changed their originally prescribed second-line regimen within 12 months after baseline (N = 713)** | | | | | | |
|---|---|---|---|---|---|---|
| Variable | Level | Viral load at 12 months < 1000 copies/ml n/N (%) | Unadjusted RR (95% CI) | P value | Adjusted RR[a] (95% CI) | P value |
| Second-line regimen | AZT/XTC/LPV/r | 284/407 (69.8) | 1 | - | 1 | - |
| | AZT/XTC/DTG | 119/137 (86.9) | 1.19 (1.07-1.33) | 0.002 | 1.20 (1.08-1.33) | 0.001 |
| | TDF/XTC/DTG | 133/169 (78.7) | 1.13 (1.02-1.25) | 0.020 | 1.11 (1.00-1.23) | 0.046 |
| Age at baseline | 15-24 | 29/51 (56.9) | 1 | - | 1 | - |
| | 25-34 | 189/257 (73.5) | 1.28 (0.98-1.65) | 0.066 | 1.31 (1.01-1.70) | 0.043 |
| | 35-44 | 216/275 (78.5) | 1.34 (1.02-1.77) | 0.035 | 1.37 (1.04-1.81) | 0.023 |
| | 45+ | 102/130 (78.5) | 1.39 (1.06-1.83) | 0.018 | 1.35 (1.02-1.79) | 0.039 |
| Gender | Male | 212/271 (78.2) | 1 | - | 1 | - |
| | Female | 324/442 (73.3) | 0.95 (0.87-1.03) | 0.23 | 0.95 (0.87-1.03) | 0.22 |
| Known tuberculosis at baseline | No | 531/703 (75.5) | 1 | - | 1 | - |
| | Yes | 5/10 (50.0) | 0.65 (0.37-1.14) | 0.13 | 0.66 (0.38-1.16) | 0.15 |
| Recent viral load (copies/ml) at baseline | 1,000 to <10,000 | 250/301 (83.1) | 1 | - | 1 | - |
| | 10,000+ | 286/412 (69.4) | 0.85 (0.79-0.92) | <0.001 | 0.84 (0.78-0.91) | <0.001 |
| Recent CD4 count (cells/μl) at baseline | ≤ 200 | 201/264 (76.1) | 1 | - | - | - |
| | 201–350 | 121/167 (72.5) | 0.96 (0.87-1.06) | 0.41 | - | - |
| | 351–500 | 79/96 (82.3) | 1.08 (0.97-1.21) | 0.16 | - | - |
| | > 500 | 59/86 (68.6) | 0.92 (0.78-1.09) | 0.33 | - | - |
| | Missing | 76/100 (76.0) | 1.01 (0.88-1.17) | 0.84 | - | - |
| Years on ART at baseline | < 2 year | 198/254 (78.0) | 1 | - | - | - |
| | ≥ 2 years | 338/459 (73.6) | 0.95 (0.88-1.01) | 0.12 | - | - |

Data are n/N (%), unless otherwise stated. [a]The primary exposure effect (viral suppression at 12 months) is adjusted for all other variables in the table as potential confounders except CD4 count and Years on ART at baseline. ART = Antiretroviral treatment, AZT = Zidovudine, DTG = Dolutegravir, EFV = Efavirenz, LVP/r = Lopinavir-ritonavir, μl = Microliter, ml = Milliliter, NVP = Nevirapine, PLHIV = People living with HIV, RR = Risk ratio, TDF = Tenofovir disoproxil fumarate, XTC = Emtricitabine or Lamivudine.
